# Assessing the causal role of sleep traits on glycated haemoglobin: a Mendelian randomization study

**DOI:** 10.1101/2020.12.18.20224733

**Authors:** Junxi Liu, Rebecca C Richmond, Jack Bowden, Ciarrah Barry, Hassan S Dashti, Iyas Daghlas, Jacqueline M Lane, Samuel E Jones, Andrew R Wood, Timothy M Frayling, Alison K Wright, Matthew J Carr, Simon G Anderson, Richard Emsley, David Ray, Michael N Weedon, Richa Saxena, Deborah A Lawlor, Martin K Rutter

## Abstract

**Objective:** To examine the effects of sleep traits on glycated haemoglobin (HbA1c).

**Design:** Observational multivariable regression (MVR), one-sample Mendelian randomization (1SMR), and two-sample summary data Mendelian randomization (2SMR).

**Setting:** UK Biobank (UKB) prospective cohort study and genome-wide association studies from the Meta-Analyses of Glucose and Insulin-related traits Consortium (MAGIC).

**Participants:** In MVR and 1SMR, participants were adults (mean (SD) age 57 (8) years; 54% female) from the UKB (n=336,999); in 2SMR, participants were adults (53 (11) years; 52% female) from MAGIC (n=46,368). All participants were adults of European ancestry.

**Exposures:** Self-reported insomnia frequency (usually *vs* sometimes or rarely/never); sleep duration: 24-hour sleep duration (hours/day); short sleep (≤6 hours *v*s 7-8 hours) and long sleep (≥9 hours *vs* 7-8 hours); daytime sleepiness and daytime napping (each consisting of 3 categories: never/rarely, sometimes, usually); chronotype (5 categories from definite morning to definite evening preference).

**Main outcome measure:** HbA1c in standard deviation (SD) units.

**Results:** Across MV, 1SMR, 2SMR, and their sensitivity analyses we found a higher frequency of insomnia (usually *vs* sometimes or rarely/never) was associated with higher HbA1c (MVR: 0.053 SD units, 95% confidence interval (0.046 to 0.061), 1SMR: 0.52, (0.42 to 0.63), 2SMR: 0.22, (0.10 to 0.35)). Results remained significant but point estimates were somewhat attenuated after excluding people with diagnosed diabetes. For other sleep traits, there was less consistency with significant associations when using some, but not all methods.

**Conclusions:** This study suggests that insomnia increases HbA1c levels. These findings could have important implications for developing and evaluating strategies that improve sleep habits to reduce hyperglycaemia and prevent diabetes.

**SUMMARY BOX:** *What is already known on this topic:*

- In observational data, insomnia, short sleep duration, and evening preference are associated with higher risk for type 2 diabetes.
- Mendelian randomization (MR) studies have not found evidence of a causal effect of short sleep on type 2 diabetes or glycaemic traits but have indicated an effect of insomnia on type 2 diabetes. It is unclear whether insomnia influences HbA1c levels, a marker of long-term hyperglycaemia, in the general population.
- Recently identified genetic variants robustly associated with insomnia, sleep duration, daytime sleepiness, napping, and chronotype can be used in MR studies to explore causal effects of these sleep traits on HbA1c levels.

*What this study adds:*

- This study suggests that a higher frequency of insomnia increases HbA1c levels in the general population and after excluding people with diabetes.
- We found no robust evidence for causal effects of other sleep traits on HbA1c levels.
- These findings improve our understanding of the impact of sleep traits on HbA1c levels and have important implications for developing and evaluating strategies that improve sleep habits to reduce hyperglycaemia and prevent diabetes.

## INTRODUCTION

Experimental studies in healthy adults have shown that interventions reducing the duration of sleep or interrupting sleep result in increased insulin resistance and higher plasma glucose levels.^1 2^ Systematic reviews and meta-analyses of prospective studies using conventional multivariable regression consistently find that both shorter^3 4^ and longer sleep duration^3^ are associated with higher risk of type 2 diabetes (T2D) (for each hour of sleep <7 hours/day there was a 9% higher T2D risk, and for each hour of sleep >7 hours/day there was a 14% higher risk^5^). Observational studies have also shown that insomnia (difficulty initiating or maintaining sleep),^6^ daytime napping,^7^ and chronotype (preference for mornings or evenings)^8^ are associated with higher T2D risk. However, causal relations are unclear from these data due to the potential for these studies to be biased by residual confounding (e.g. from physical activity and diet) and reverse causality (e.g. from nocturia and neuropathic pain).

Mendelian randomization (MR) analysis, which uses genetic variants as instrumental variables to appraise causal effects of exposures (e.g. sleep traits) on outcomes (e.g. glycaemic levels), is less prone to confounding by socioeconomic and behavioural factors, or to reverse causality, than conventional observational multivariable regression (MVR).^9^ As MR has different key sources of bias (e.g. horizontal pleiotropy) to MVR, where there is consistency between the two methods this increases confidence in there being a causal effect.^10^ Three recently published studies,^11-13^ using two-sample Mendelian randomization (2SMR), find no evidence of causal effects of sleep duration on T2D and/or related glycaemic traits – apart from one that suggested that longer sleep duration could potentially increase insulin production.^12^ Prior MR studies have suggested that insomnia might have a causal role in increasing T2D risk,^14 15^ but they did not assess whether insomnia influences glycaemic levels in the general population. These prior studies may have been limited by low statistical power and suffered from weak instrument bias.^16^ Understanding the impact of sleep traits on glycaemic levels in the general population would have profound public health implications for the prevention of diabetes.

Our primary aim was to explore effects of sleep traits (i.e., insomnia,^14^ sleep duration,^17^ daytime sleepiness,^18^ daytime napping,^19^ and chronotype^20^) on average glycaemic levels assessed by HbA1c in general population. Our secondary aim was to assess the effects of these sleep traits on non-fasting glucose levels. We extend prior work by exploring the effects of sleep traits on HbA1c and glucose levels using several complementary approaches including traditional MVR and novel MR methods within the UK Biobank (UKB; n=336,999), and using genome-wide association studies from the Meta-Analyses of Glucose and Insulin-related traits Consortium (MAGIC) (n=46,368).

## MATERIALS AND METHODS

### Overview

We assessed relations between sleep traits and measures of glycaemia using traditional observational epidemiology – cross-sectional observed confounder-adjusted MVR; one-sample MR (1SMR; in which the relationships between sleep trait genetic variants and our study exposures and outcomes were assessed in *the same study*) and 2SMR (in which the genetic variant-exposures and genetic variant-outcome associations were assessed in two *different studies*). Importantly, we undertook extensive sensitivity analyses, including novel methods assessing the robustness of 1SMR to bias by unbalanced horizontal pleiotropy, weak instruments, and winner’s curse.^21^

### The UK Biobank

For our confounder-adjusted MVR and 1SMR analyses, we used data on UKB participants. Between 2006 and 2010, UKB recruited 503,317 participants aged 40-69 years, from 9.2 million adults who were invited to take part (5.5% response rate).^22 23^ All participants provided informed consent. Baseline data on self-reported sleep traits and other lifestyle and socio-demographic characteristics were obtained using a touchscreen questionnaire. At the same time, anthropometric measures and non-fasting venous blood samples were taken. These samples were used for HbA1c, non-fasting glucose, and chip based genome-wide analyses; the latter providing single-nucleotide polymorphism (SNP) data. HbA1c (mmol/mol) was measured in red blood cells by HPLC on a Bio-Rad VARIANT II Turbo analyzer and non-fasting glucose (mmol/l) was assayed in serum by hexokinase analysis on a Beckman Coulter AU5800.^24^

UKB included 488,377 successfully genotyped participants: 49,979 using the UK BiLEVE chip and 438,398 using the UKB axiom chip. Pre-imputation quality control, phasing and imputation of the UKB genetic data have been described.^25^ Of the 488,377 successfully genotyped participants, 488,288 were with available phenotypic data, among which, 409,629 were self-reported as “White British”. Of these 409,629, 72,617 were excluded, after accounting for duplication, based on sex mismatch (n=312), sex chromosome aneuploidy (n=556), outliers in heterozygosity and missing rates (n=731), and relatedness based on estimated kinship coefficients (n=71,274).^26^ Additionally, 13 participants who had withdrawn consent prior to our analyses were removed (by 04 February 2020). Thus, 336,999 participants were included in the final analysis (**Figure 1**).

**Figure 1.**
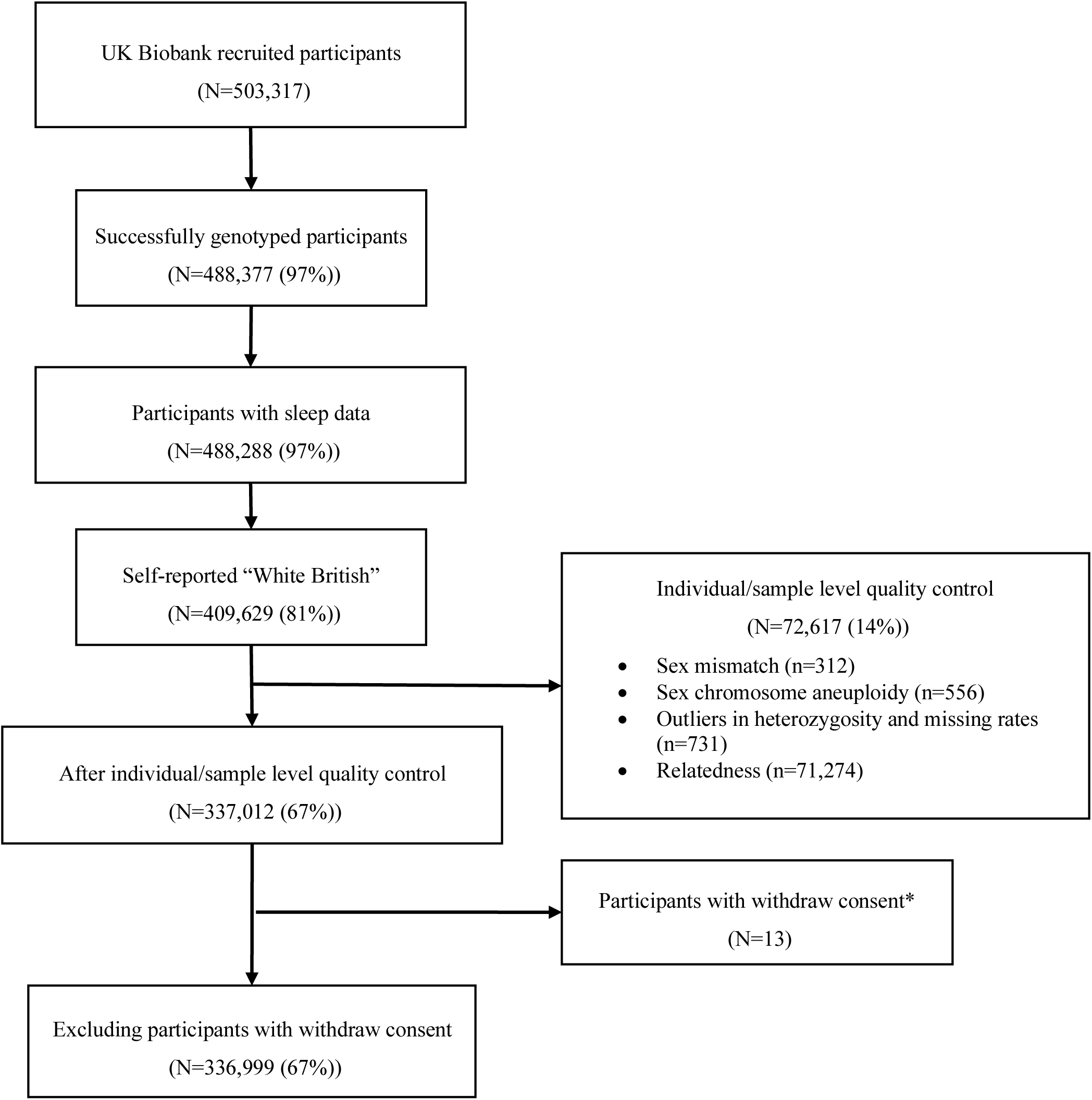
Flowchart of the participants included in final analyses of the UK Biobank. % values in brackets are of the total recruited to UK Biobank Participants with withdraw consent by 04 February 2020

We assessed associations between HbA1c/glucose (outcomes) and seven self-reported sleep traits (exposures): insomnia frequency categorised as usually *vs* sometimes or rarely/never; total 24-hour sleep duration (reported in whole hours), short sleep (≤6 hours compared to 7 or 8 hours) and long sleep duration (≥9 hours compared to 7 or 8 hours). Responses to questions on daytime sleepiness and daytime napping were categorized into three frequency levels (“Never/rarely”, “Sometimes”, and “Usually”). Chronotype was coded into five categories (“Definitely a ‘morning’ person”, “More a ‘morning’ than ‘evening’ person”, “Do not know”, “More an ‘evening’ than a ‘morning’ person”, and “Definitely an ‘evening’ person”), meaning that our results reflect a difference in mean HbA1c/glucose *per category increase* towards evening preference. Those who responded “Do not know” or “Prefer not to say” were treated as missing data for all sleep traits, except for chronotype where “Do not know” was treated as an intermediate category (details in **Supplementary Methods**).

### Meta-Analyses of Glucose and Insulin-related traits Consortium

For 2SMR analyses, sex-combined meta-analysis summary statistics of genetic variants related to HbA1c (n=46,368, mean (SD) age 53 (11) years, 52% female), from 23 genome-wide association studies (GWAS))^27^ and fasting glucose (n=46,186 mean (SD) age, 52 (13) years 56% female) from 21 GWAS)^28^ were obtained from the MAGIC consortium. Participants were of European descent without diagnosed diabetes.

### Genetic variants – instrumental variables

In 1SMR and 2SMR analyses, we used genetic variants identified in the GWAS of the seven self-reported sleep traits as passing the GWAS multiple testing p-value threshold (<5*10^−8^).^14 17-20^ They were described in **Supplementary Table 1**.

### Statistical analyses

In all three analyses (MVR, 1SMR, and 2SMR), we coded the effect of sleep exposures in terms of a unit increase or a category increase in each trait on the outcome, except for insomnia, short sleep duration and long sleep duration, which we used as binary exposure variables. For insomnia frequency, we compared people reporting “Usually” *vs* those reporting “Sometimes” or “Rarely/Never”; for short sleep, we compared people reporting sleep duration ≤6 hours with those with normal duration (7 or 8 hours); for long sleep, we compared people reporting sleep duration ≥9 hours with those with normal duration (7 or 8 hours). For daytime sleepiness and daytime napping, the category increase reflects more frequent sleepiness and napping, respectively. For chronotype, the category increase reflects a tendency to greater evening preference.

HbA1c data were right skewed in UKB and the units used to measure HbA1c differed between UKB (mmol/mol) and the cohorts contributing to MAGIC (%). Therefore, we natural log-transformed the HbA1c levels in UKB and then converted them into SD units (HbA1c: 1 SD was equal to 0.15 log mmol/mol). For 2SMR, we also presented results in SD units of the summary data from MAGIC (1SD was equal to an HbA1c value of 0.53%, ≈ 6 mmol/mol). Thus, for all analyses (MVR, 1SMR, and 2SMR), we estimated the mean difference in HbA1c SD per 1 unit or category increase in the sleep traits (i.e., 24-hour sleep duration, daytime sleepiness, daytime napping, and chronotype) except for insomnia, short sleep (≤6 hours *vs* 7-8 hours), and long sleep (≥9 hours *vs* 7-8 hours). For these binary exposures, in MVR and 1SMR we estimated the average difference in HbA1c under the counterfactual assumption which provides an estimate of the difference between everyone (in the population of interest) experiencing the exposure (i.e. assuming exposure prevalence is 100%) compared to no-one experiencing the exposure (assuming exposure prevalence: 0%).^29^ To enable the comparison of the 2SMR estimates to the MVR and 1SMR results, we converted the results of the SNP-binary sleep trait from the multiplicative log odds scale to a difference in risk scale by *β* = log *OR* * µ * (1-*µ*), *se* = *se*_log *OR*_ * µ * (1-*µ*), with *µ* = *n*_*case*_ /(*n*_*case*_ + *n*_*control*_).^30^

#### Covariates in multivariable-adjusted regression models (observational analysis)

We considered the following potential confounders of the observational associations (MVR analyses) between sleep traits and glycaemic levels: baseline age, sex, smoking, alcohol intake, Townsend residential area deprivation score, university education, vigorous physical activity levels, and body mass index (BMI). BMI was derived from weight and height measured at UKB assessment centres. Deprivation scores were derived from residential postcodes (wards for Scotland).^31^ Data on all other confounders were taken from baseline questionnaire responses (details in **Supplementary Methods**). We also adjusted for population stratification in the MVR analyses by including the top 40 genetic principal components of ancestry and UKB assessment centre. We ran two MVR models: In model 1, the main model, we adjusted for: age, sex, smoking, alcohol intake, Townsend area deprivation index, university education, and vigorous physical activity, top 40 genetic principal components and assessment centre (referred to as ‘MVR’ in Results figures). In model 2, a supplementary model, we additionally adjusted for BMI due to uncertainty as to whether BMI was a confounder or a mediator on the causal pathway between sleep traits and glycaemic levels.

#### One-sample Mendelian Randomization

For 1SMR, the genetic variants were extracted from the UKB Haplotype Reference Consortium reference panel dataset. These data have undergone extensive quality control checks including removal of related participants (third degree or closer) and non-White British participants based on questionnaire and PCA^32^ (details in **Supplementary Methods**). Given the genetic variants of sleep traits were predominantly identified in the UKB (**Supplementary Table 1**), to account for weak instrument bias, we applied unweighted allele scores rather than weighted allele scores.^33^ Unweighted allele scores were generated as the total number of adverse sleep trait increasing alleles present for each participant (evening preference alleles for chronotype). Two-stage least squares (2SLS) instrumental variable analyses were performed with adjustment for assessment centre and 40 genetic principal components to minimize confounding by population stratification, as well as baseline age, sex and genotyping chip to account for known confounders and to reduce random variation (referred to as ‘1SMRmain’ in Results figures).

#### Two-sample Mendelian Randomization

We conducted 2SMR analyses of sleep traits with glycaemic measures using the summary associations between the genetic instruments and sleep traits identified in the respective GWAS^14 17-19^ (sample 1) (**Supplementary Table 1**) and estimates of the associations between the genetic instruments and glycaemic measures (HbA1c and fasting glucose)^27 28^ from MAGIC (sample 2). Analyses were conducted using the “TwoSampleMR” package in R (version MRCIEU/TwoSampleMR@0.4.26).^34^ If a SNP was unavailable in the outcome GWAS summary statistics, we identified a proxy in strong linkage disequilibrium (LD) with the missing SNP (r^2^>0.8). The effect allele frequency of the outcome summary data was misinterpreted in the “TwoSampleMR” package. Therefore, we manually corrected the effect allele frequency of all the merged palindromic SNPs according to the data (i.e., minor allele frequency) downloaded from the MAGIC and the information of minor allele obtained from the ensembl (https://www.ensembl.org/index.html). We then performed harmonization of the direction of effects between SNPs in the exposure and outcome GWAS. Palindromic SNPs were harmonized if they were aligned and the minor allele frequency was <0.3, otherwise they were excluded. In the primary analysis, we used the inverse-variance weighted (IVW) regression under a multiplicative random-effects model^35^ (weights are equal to the inverse of variance of SNP-outcome associations; referred to as ‘2SMRmain’ in Results figures), to obtain causal effects of sleep traits on HbA1c and fasting glucose. Estimates were compared with those obtained from the 1SMR analysis.

### Sensitivity analyses

To account for the potential impact of treatment for diabetes (either by lifestyle intervention or insulin) and the effect of diabetes on HbA1c or glucose levels, we repeated MVR and main 1SMR analyses in UKB participants after excluding those reporting a doctor diagnosis of diabetes, being treated with insulin and those with a baseline HbA1c ≥ 48 mmol/mol (equivalent to ≥ 6.5%, the threshold for diagnosing diabetes). Information on self-reported diabetes and insulin medication were obtained from the baseline questionnaire.

#### Assessing Mendelian randomization assumptions and evaluating bias

Mendelian randomization analysis requires various assumptions to be satisfied in order to estimate causal effects: that the genetic instrument is robustly associated with the exposure (instrument strength); that the genetic instrument is independent of potential confounding factors of the exposure-outcome association (no confounding); and that the genetic instrument influences the outcome exclusively through its effect on the exposure (no horizontal pleiotropy). Both 1SMR and 2SMR analysis are vulnerable to bias if these assumptions are violated; they can also be biased by winner’s curse (see below). Various steps were taken to assess the MR assumptions and to evaluate bias, as outlined below.

Instrument strength was investigated using the first-stage F-statistic and r^2^ values in both 1SMR and 2SMR. In 1SMR main analysis, we examined associations between the allele scores and variables potentially related to HbA1c (smoking, alcohol, deprivation, university education, vigorous physical activity, and BMI) to explore violation of the no confounding and no horizontal pleiotropy assumptions by specific known risk factors for variation in HbA1c that were observed in UKB. To assess potential bias due to unbalanced horizontal pleiotropy, we also explored between-SNP heterogeneity using the Sargan test.^36^ To further explore this potential bias in our 1SMR, we applied a novel method that allows bias introduced by weak instruments (which would bias our results towards the confounded results),^16^ and unbalanced horizontal pleiotropy, to be addressed in a 1-sample setting using 2SMR approaches.^21^ This method allowed us to report IVW, MR-Egger,^37^ and least absolute deviation (LAD) regression (similar to the weighted median (WM))^38^ in our 1SMR analyses. In the figures presented in the Results, we referred to these as 1SMRsensitivity1, 1SMRsensitivity2, and 1SMRsensitivity3, respectively. Full details of these novel methods were provided in **Supplementary Methods**.

Winner’s curse can occur when the study in which the genetic variants were identified as genome-wide significance (p-value<5×10^−8^) (our method for selecting genetic instruments) is the same study used to perform MR analysis.^39^ Bias introduced by winner’s curse was likely in our 1SMR, because the genetic variants for the sleep traits were predominantly identified in UKB. We would expect this to bias results towards the null. To address this, we identified subsets of genome-wide significance SNPs of some sleep traits (i.e., 108 for insomnia,^14^ 19 for daytime sleepiness,^18^ 17 napping,^19^ and 72 for chronotype^20^) in other independent GWAS that did not include UKB. Details of these subsets were provided in **Supplementary Methods**. The novel 1SMR weak and pleiotropy-robust methods were then applied to use these SNP subsets. This strategy removes winner’s curse and is weak-instrument robust.

In 2SMR, to explore directional horizontal pleiotropy, we compared results from the IVW regression with two pleiotropy-robust MR methods: MR-Egger^37^ and WM^38^ (in Results figures, we referred to these as 2SMRsensitivity1 and 2SMRsensitivity2, respectively).

#### Additional analysis

To rule out the potential reverse causality of an effect of any sleep traits on HbA1c, we would conduct bidirectional MR analyses (1SMR and 2SMR) to assess the potential inverse association. Full details of the bidirectional MR were provided in **Supplementary Methods**.

As noted above, it is unclear whether BMI is a confounder or mediator of the sleep traits-HbA1c association. Given BMI is strongly associated with HbA1c, we agreed that in the context of any MR data supporting for effects of sleep traits on HbA1c, if the gene allele score was related to BMI, we would further explore this in multivariable MR (MVMR).^40^ Full details of MVMR were provided in **Supplementary Methods**.

### Patient and public involvement

Two patients with T2D, under the care of MKR, helped develop the original idea for this research and these discussions highlighted the importance of potential reverse causality and confounding in MVR analyses, and hence the need to explore the research questions with additional methods as mentioned above. Participants in UK Biobank are regularly updated about research undertaken in the study through newsletters and invitations to meetings where scientists using UKB data present their results. The results of this research will be similarly disseminated to study participants, relevant stakeholders and the broader public as appropriate.

## RESULTS

### Baseline characteristics

In the 336,999 participants included in this study, the mean (SD) age was 56.9 (8.0) years and 54% were female **(Table 1)**. There were 5% of participants with a self-reported diagnosis of diabetes, 0.4% reported using insulin, and 3% with a HbA1c value ≥ 48 mmol/mol. The frequency of insomnia was reported to be ‘usually’ by 28% of the cohort; mean sleep duration was 7.2 (1.1) hours; daytime sleepiness was reported as ‘often/always’ by 3% and ‘sometimes’ by 20%; daytime napping was reported as ‘often’ by 5% and ‘sometimes’ by 38%; definite morning chronotype was reported by 24% and definite evening chronotype by 8% of participants. The median (interquartile range) HbA1c was 35 (33, 37) mmol/mol in people without diabetes and 50 (43, 59) mmol/mol in those with diabetes; corresponding non-fasting glucose levels were 4.9 (4.6, 5.3) mmol/L and 6.5 (5.3, 8.9) mmol/L respectively.

**Table 1.**
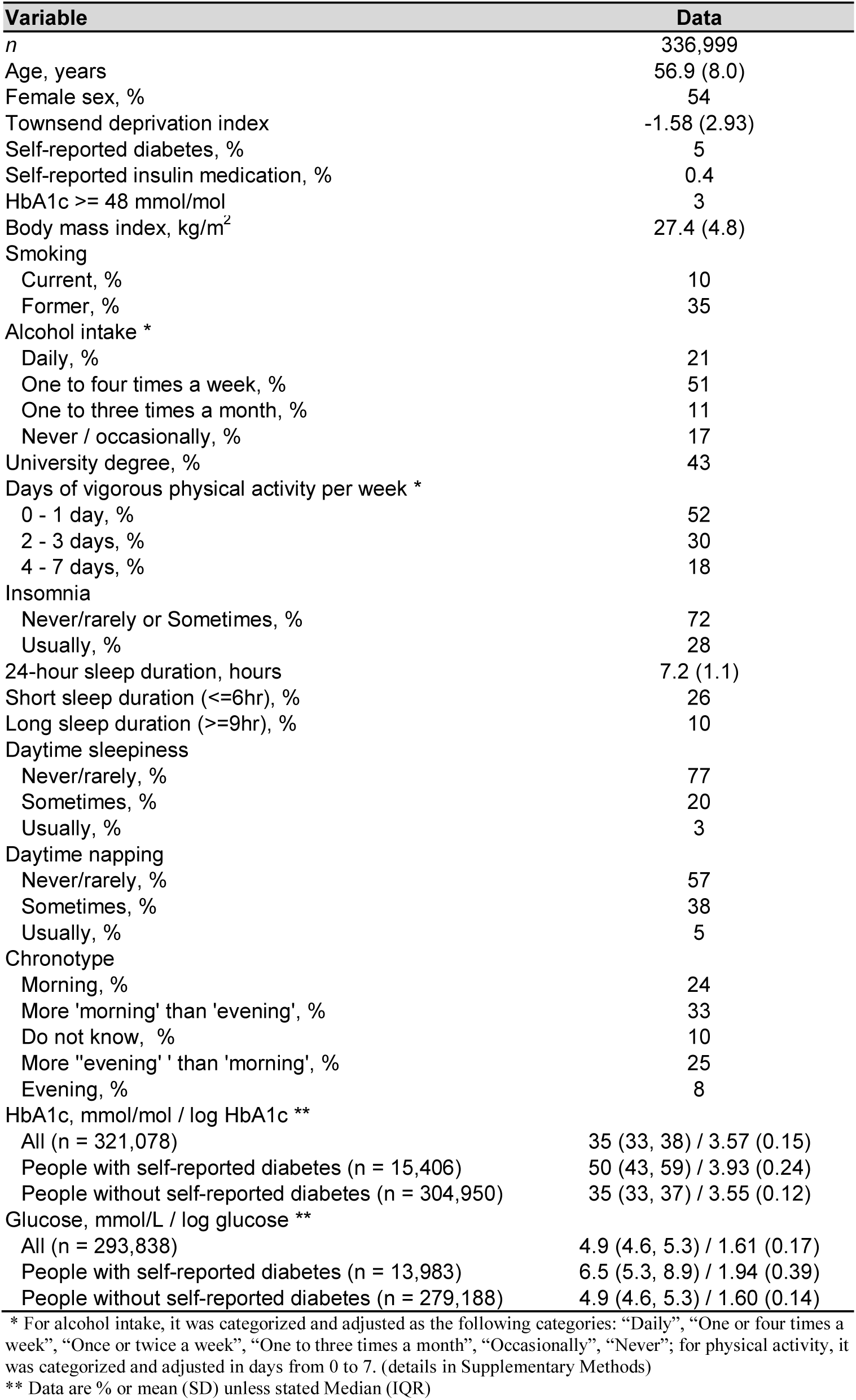
Baseline characteristics of the participants included in final analyses of the UK Biobank

People who participated in this study (n=336,999) were older and were less likely to be female when compared to those who were excluded in the whole UKB cohort (n=165,631) (age 56.9 vs. 55.8 years; 54% vs. 56% female). Included participants were also more highly educated, less deprived, and less likely to smoke. HbA1c, glucose, other sleep traits, BMI, alcohol consumption, and physical activity were similar between the included and excluded participants (**Supplementary Table 2)**.

### Associations of sleep traits with HbA1c and glucose levels

There was consistent evidence across all analyses that more frequent insomnia (usually *vs* sometimes or rarely/never) resulted in higher HbA1c levels (MVR: 0.053 SD units, 95% confidence interval (0.046 to 0.061), main 1SMR (0.52, (0.42 to 0.63)), and main 2SMR (0.22, (0.10 to 0.35); **Figure 2**). With exclusion of ∼ 15,000 (4%) participants with self-reported diabetes or using insulin, results of MVR and 1SMR were attenuated towards the null but remained consistent with main analyses (**Supplementary Table 3**). After an additional exclusion of those with HbA1c ≥ 48 mmol/mol (additional ∼ 2000 participants), results were essentially the same as those only excluding participants with diabetes/using insulin (**Supplementary Table 3**). Results of sensitivity analyses accounting for weak instrument bias, unbalanced horizontal pleiotropy, and winner’s curse were consistent with the main 1SMR and 2SMR results (**Supplementary Table 3**). When we adjusted for BMI in MVMR of the effect of insomnia on HbA1c, the results were consistent with the main analyses (**Supplementary table 4**). More frequent insomnia symptoms (usually *vs* sometimes or rarely/never) was associated with higher non-fasting glucose levels in the MVR and main 1SMR analyses, whereas, the positive associations were weaker in magnitude and consistent with the null in 1SMR pleiotropy-robust sensitivity and in all 2SMR analyses (**Supplementary Table 5**).

**Figure 2.**
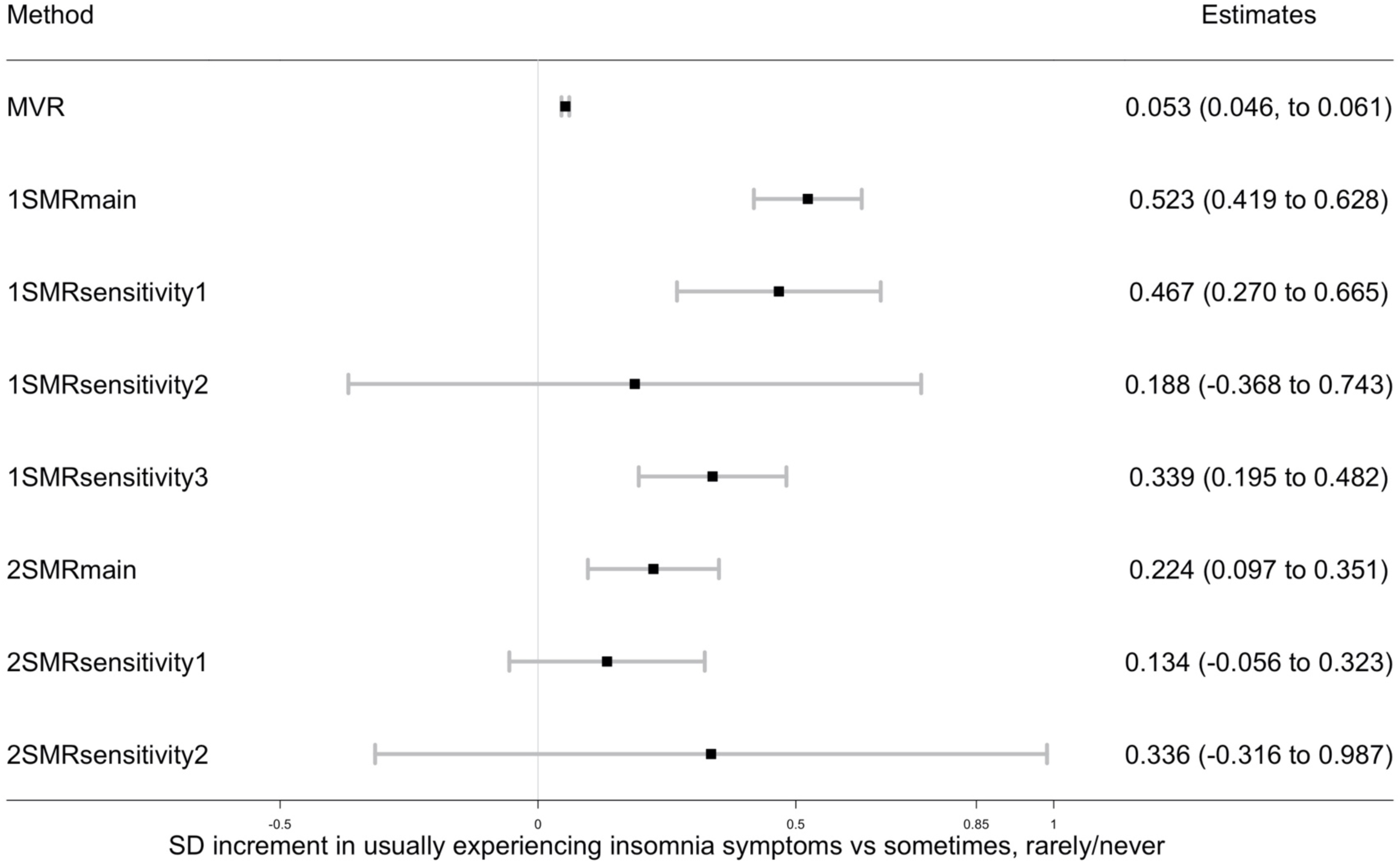
Associations of insomnia with HbA1c in observational multivariable regression analysis, one-sample Mendelian randomization in the UK Biobank and two-sample Mendelian randomization in MAGIC. Data are SD increment (95% CI) in HbA1c comparing people usually experiencing insomnia vs sometimes, really/never MVR: multivariable regression adjusted for sex, age, assessment centre, 40 genetic principal components, smoking, alcohol intake, Townsend deprivation, education, and physical activity 1SMRmain, 1SMRsensitivity1, 1SMRsensitivity2, and 1SMRsensitivity3 were equivalent to two-stage least square, IVW, MR-Egger, and least absolute deviations (LAD) regression in one-sample Mendelian randomization respectively. 2SMRmain, 2SMRsensitivity1, 2SMRsensitivity2 were equivalent to IVW, Weighted Median, and MR-Egger in two-sample Mendelian randomization respectively 1SD HbA1c in the UK Biobank is 0.15 log mmol/mol; 1SD HbA1c in the MAGIC is 0.53% (∼6 mmol/mol)

Longer 24-hour sleep duration, treated as a continuous variable, was associated with lower HbA1c levels in the MVR (−0.007 (95% CI: −0.010 to −0.003) SD per 1 hour longer) and the main 1SMR analysis (−0.11 (−0.15 to −0.07)), but were close to the null in the 1SMR pleiotropy-robust sensitivity analyses. 2SMR analyses did not support a strong causal evidence (**Figure 3** and **Supplementary Table 3**). The inverse MVR and 1SMRmain estimates remained after excluding participants with self-reported diabetes or insulin use and those with HbA1c ≥ 48 mmol/mol (**Supplementary Table 3**).

**Figure 3.**
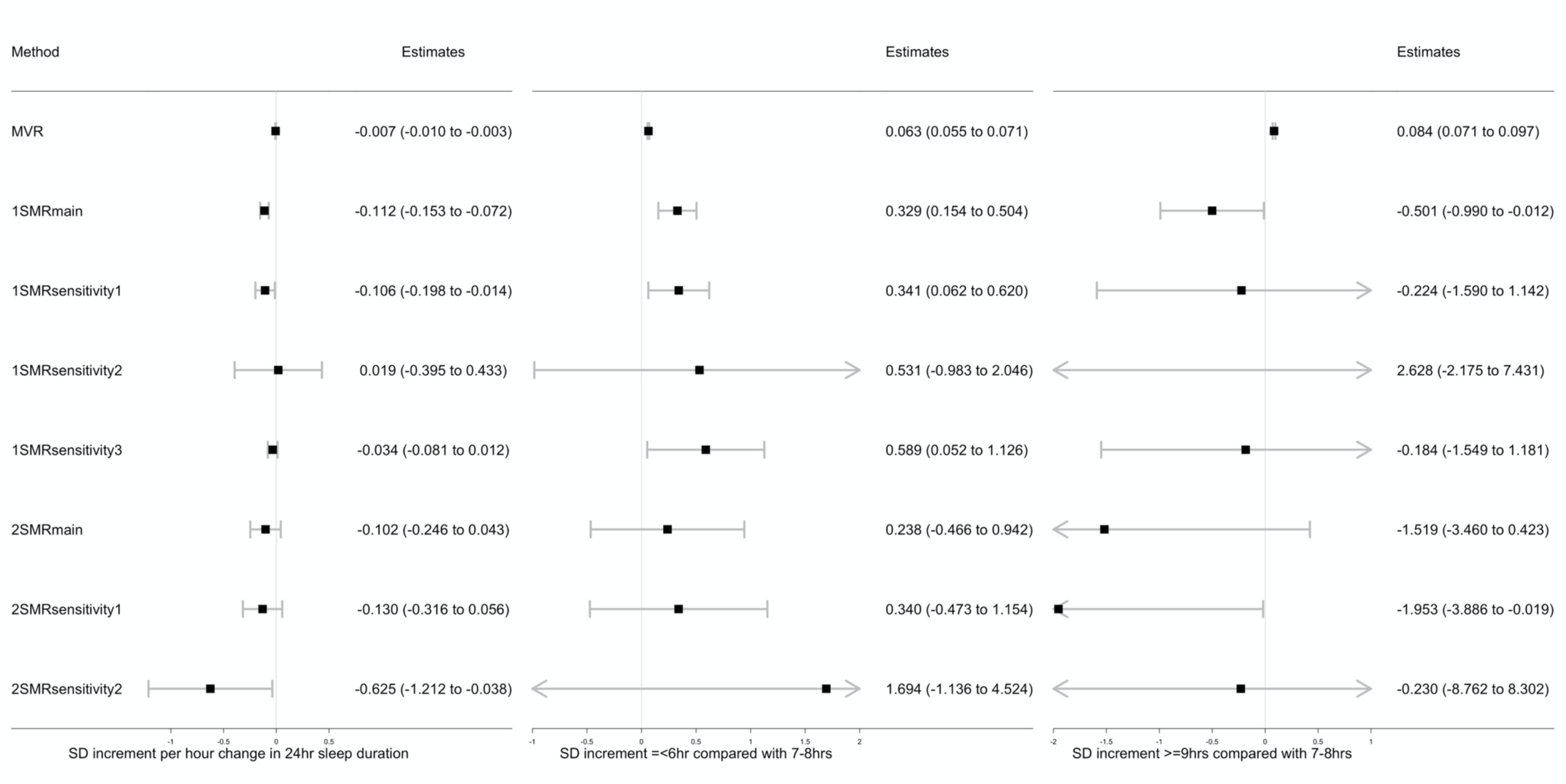
Associations of sleep duration traits with HbA1c in observational multivariable regression analysis, one-sample Mendelian randomization in the UK Biobank and two-sample Mendelian randomization in the MAGIC. Data are SD increment (95% CI) in HbA1c in relation to differences in sleep duration MVR: multivariable regression adjusted for sex, age, assessment centre, 40 genetic principal components, smoking, alcohol intake, Townsend deprivation, education, and physical activity 1SMRmain, 1SMRsensitivity1, 1SMRsensitivity2, and 1SMRsensitivity3 were equivalent to two-stage least square, IVW, MR-Egger, and least absolute deviations (LAD) regression in one-sample Mendelian randomization respectively 2SMRmain, 2SMRsensitivity1, 2SMRsensitivity2 were equivalent to IVW, Weighted Median, and MR-Egger in two-sample Mendelian randomization respectively 1SD HbA1c in the UK Biobank is 0.15 log mmol/mol; 1SD HbA1c in the MAGIC is 0.53% (∼6 mmol/mol)

In the observational MVR analysis, both short (≤6 hours) and long sleep duration (≥9 hours), treated as binary variables, were associated with higher HbA1c when compared to having a normal 7-8 hours per day sleep duration; (short sleep: 0.063, (0.055 to 0.071); long sleep: 0.08, (0.07 to 0.10)). However, in the main 1SMR analysis, there was evidence that short sleep duration increased HbA1c levels (0.33, (0.15 to 0.50)) but that long sleep duration reduced HbA1c (−0.50, (−0.99 to −0.01)). For short sleep duration, 1SMR pleiotropy-robust sensitivity analyses showed that the direction and strength of these associations were similar but there were wide confidence intervals around point estimates. For long sleep duration, 1SMR sensitivity analyses showed similar directions and strengths of association in only 2 out of 3 analyses and there were wide confidence intervals around all point estimates. The main 2SMR analysis and the related pleiotropy-robust sensitivity analyses did not support strong associations of short sleep duration or long sleep duration with HbA1c (**Figure 3** and **Supplementary Table 3**). Excluding participants with self-reported diabetes or insulin use and those with HbA1c ≥ 48 mmol/mol, the positive associations of short sleep with HbA1c in MVR and 1SMR remained but the associations of long sleep with lower HbA1c were attenuated to the null (**Supplementary Table 3**). Taking non-fasting glucose as the outcome, longer 24-hour sleep duration, short and long sleep were all positively associated with glucose level in MVR, whereas 1SMR, 2SMR, and all their sensitivity analyses did not consistently support causal effects (**Supplementary Table 5**).

MVR and main 1SMR showed that more frequent daytime sleepiness (MVR: 0.09, (0.08 to 0.10); 1SMRmain: 0.14, (0.02 to 0.27)) and daytime napping (MVR: 0.09, (0.08 to 0.10); 1SMRmain: 0.09, (0.03 to 0.14)) were associated with higher HbA1c levels, though 1SMR sensitivity analyses, including those exploring winner’s curse and weak instrument bias, and all 2SMR analyses, were not supportive of causal effects (**Figure 4** and **Supplementary Table 3**). Excluding participants with diabetes or insulin treatment and those with HbA1c ≥ 48 mmol/mol, the MVR estimates remained but the main 1SMR estimates were attenuated towards the null (**Supplementary Table 3**). There was also limited evidence of a causal effect of daytime sleepiness and daytime napping on glucose (**Supplementary Table 5**).

**Figure 4.**
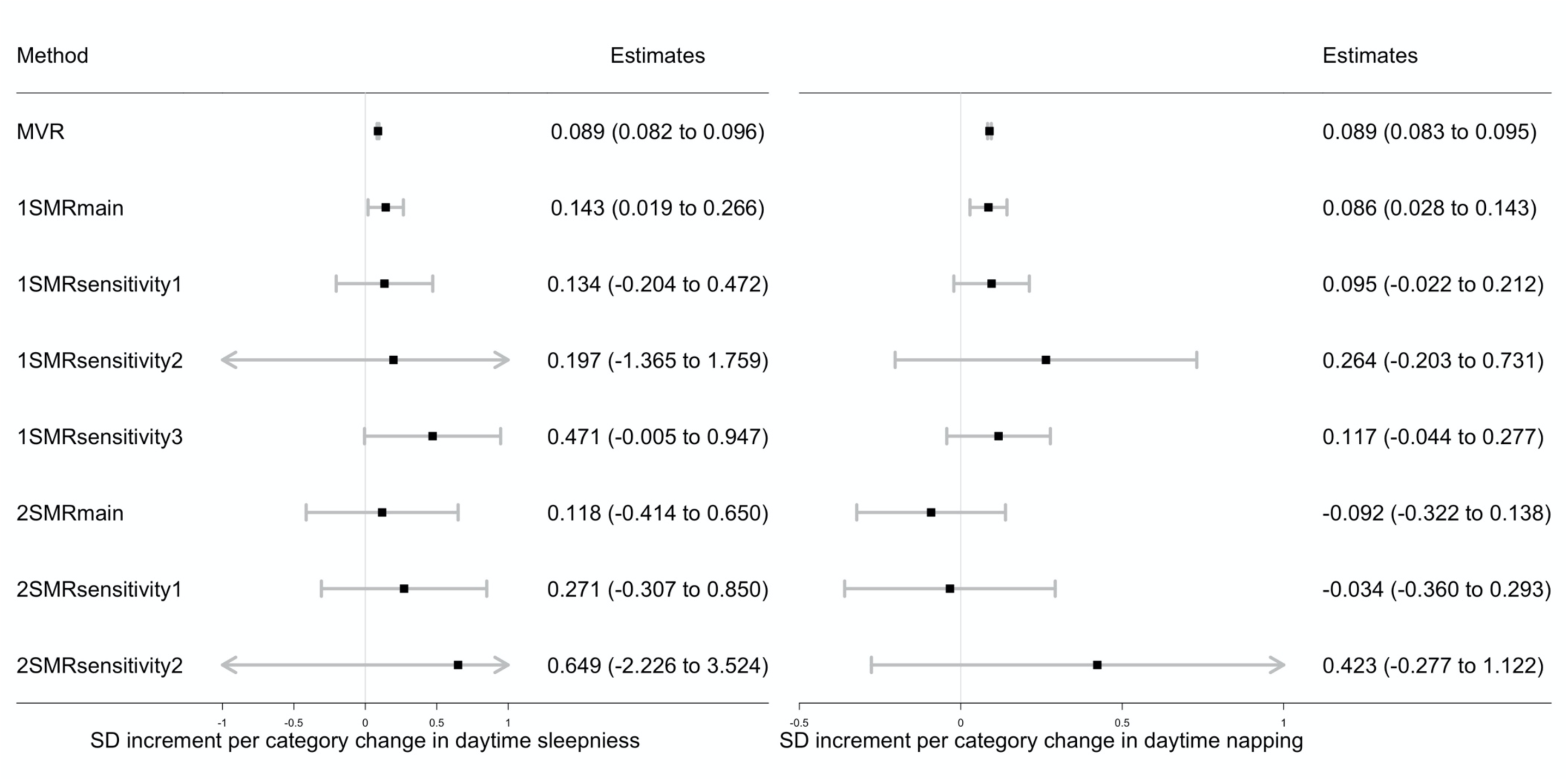
Associations of daytime sleepiness and daytime napping with HbA1c in observational multivariable regression analysis, one-sample Mendelian randomization in the UK Biobank and two-sample Mendelian randomization in the MAGIC. Data are SD increment (95% CI) in HbA1c in relation to higher frequencies of daytime sleepiness or daytime napping MVR: multivariable regression adjusted for sex, age, assessment centre, 40 genetic principal components, smoking, alcohol intake, Townsend deprivation, education, and physical activity 1SMRmain, 1SMRsensitivity1, 1SMRsensitivity2, and 1SMRsensitivity3 were equivalent to two-stage least square, IVW, MR-Egger, and least absolute deviations (LAD) regression in one-sample Mendelian randomization respectively 2SMRmain, 2SMRsensitivity1, 2SMRsensitivity2 were equivalent to IVW, Weighted Median, and MR-Egger in two-sample Mendelian randomization respectively 1SD HbA1c in the UK Biobank is 0.15 log mmol/mol; 1SD HbA1c in the MAGIC is 0.53% (∼6 mmol/mol)

Evening preference was associated with higher HbA1c levels in MVR (0.008, (0.006 to 0.011)) and main 1SMR (0.022, (0.004 to 0.039)). However, these results were attenuated and included the null in all 1SMR pleiotropy-robust sensitivity analyses (including accounting for winner’s curse) and in all 2SMR analyses (**Figure 5** and **Supplementary Table 3**). Both MVR and main 1SMR estimates included the null when excluding participants with diabetes or insulin treatment and those with HbA1c ≥ 48 mmol/mol (**Supplementary Table 3**). Results for glucose were largely consistent with findings for HbA1c (**Supplementary Table 5**).

**Figure 5.**
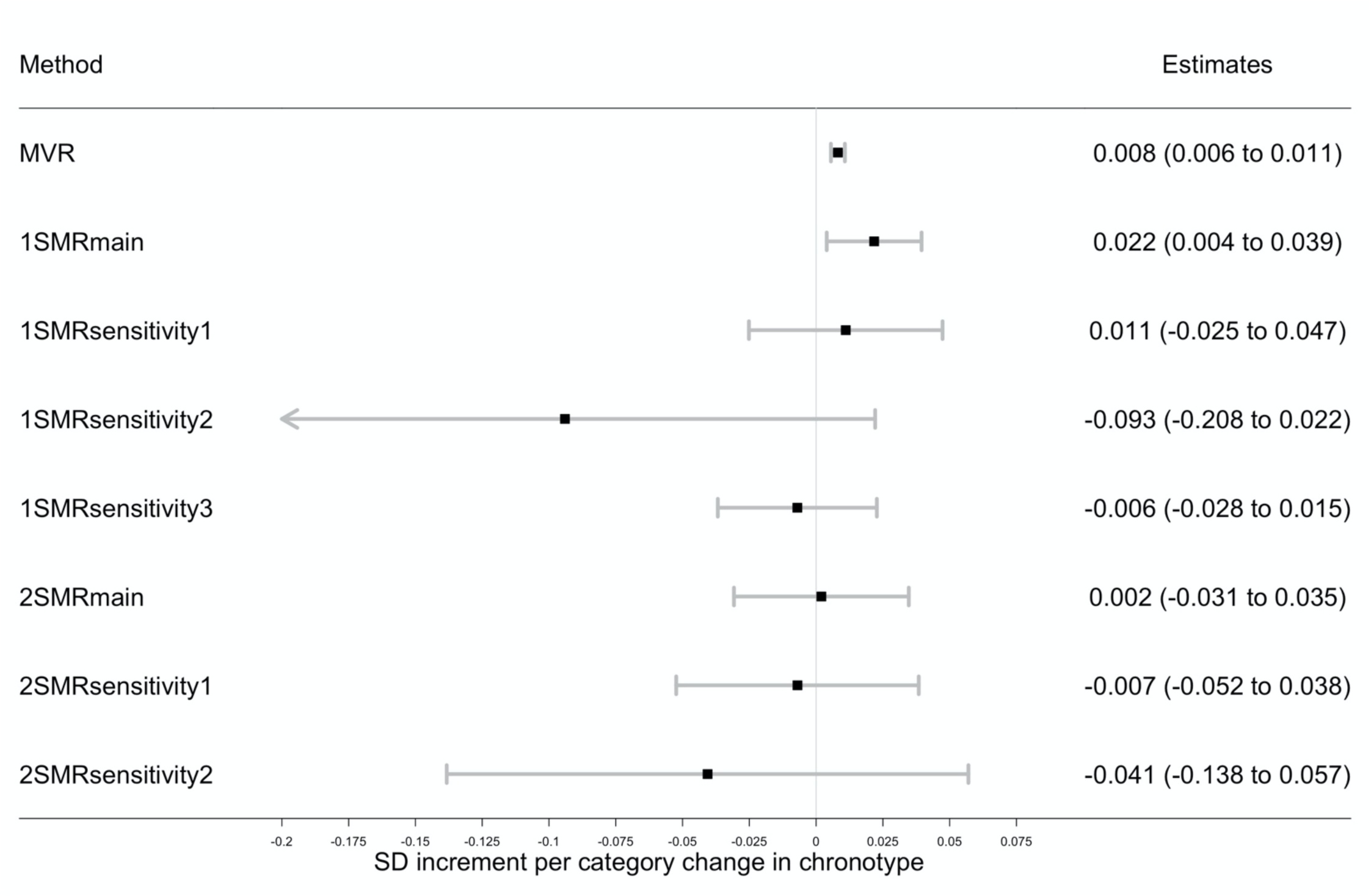
Associations of chronotype (evening preference) with HbA1c in observational multivariable regression analysis, one-sample Mendelian randomization in the UK Biobank and two-sample Mendelian randomization in the MAGIC. Data are SD increment (95% CI) in HbA1c in relation to having a higher category of evening preference MVR: multivariable regression adjusted for sex, age, assessment centre, 40 genetic principal components, smoking, alcohol intake, Townsend deprivation, education, and physical activity 1SMRmain, 1SMRsensitivity1, 1SMRsensitivity2, and 1SMRsensitivity3 were equivalent to two-stage least square, IVW, MR-Egger, and least absolute deviations (LAD) regression in one-sample Mendelian randomization respectively 2SMRmain, 2SMRsensitivity1, 2SMRsensitivity2 were equivalent to IVW, Weighted Median, and MR-Egger in two-sample Mendelian randomization respectively 1SD HbA1c in the UK Biobank is 0.15 log mmol/mol; 1SD HbA1c in the MAGIC is 0.53% (∼6 mmol/mol)

Most of the MVR estimates of sleep traits with HbA1c and glucose were attenuated to some extent after adjusting for BMI (**Supplementary Table 3** and **Supplementary Table 5**)

### Testing Mendelian randomization assumptions

In 1SMR, the variation in sleep traits explained by the allele scores varied from 2.14% (F-statistic 7359) for chronotype to 0.07% (F-statistic 181) for long sleep duration (**Supplementary Table 1**). The mean F-statistics of individual SNPs for each of the sleep traits calculated in UKB were ≥ 23, except for insomnia for which it was 8 (**Supplementary Table 6**). In 2SMR, the variance explained by the combined SNPs was also highest for chronotype (2.09%, mean F-statistic 60) and lowest for long sleep duration (0.06%, mean F-statistic 41) (**Supplementary Table 1**). Details of any proxy SNPs used in the 2SMR are described in **Supplementary Table 7**.

After accounting for multiple testing (p<0.05/6 = 0.008), the allele score for insomnia was associated with six potential risk factors for variation in glycaemic traits that might result in directional pleiotropy (smoking, alcohol, Townsend residential deprivation index, BMI, and university education). To differing degrees, the allele scores of short sleep duration, daytime sleepiness, daytime napping, and chronotype also associated with one or more of these risk factors, whereas allele scores of total sleep duration and long sleep duration were not substantively associated with these risk factors (**Supplementary Table 8**).

In 1SMR, the Sargan test indicated heterogeneity of the SNP estimates for all sleep traits with HbA1c and glucose, which could suggest horizontal pleiotropy. However, MR-Egger intercepts did not provide evidence of any directional pleiotropy (**Supplementary Table 3** and **Supplementary Table 5**).

## DISCUSSION

Across MVR, 1SMR, 2SMR, and their sensitivity analyses, we found consistent evidence that insomnia increases HbA1c levels. Whereas, evidence supporting causal effects of other sleep traits on HbA1c and glucose levels was less clear.

### Comparison with other studies

Previous observational studies have shown that insomnia,^6^ short and long sleep duration,^41 42^ daytime sleepiness,^43^ daytime napping,^44^ and evening preference^45^ are associated with hyperglycaemia or higher T2D risk. Our findings support previous observational^6^ and MR^14 15^ studies showing that insomnia is associated with higher T2D risks. Here, we extended these findings by showing an effect of insomnia on HbA1c in the wider population and after excluding people with diabetes. The finding that insomnia was more strongly linked to hyperglycaemia than other sleep traits is in-keeping with a suggestion made previously based on observational data.^46^

With regards to 24-hour sleep duration, short and long sleep, prior 2SMR studies^11-13^ found limited evidence of causal effects of these exposures on HbA1c, glucose, or T2D. By comparing estimates of 2SMR with MVR associations and 1SMR analyses, and including novel 1SMR sensitivity analyses (i.e., weak instrument and pleiotropy-robust methods in 1SMR),^21^ our study considerably strengthens previous null findings. Our results together with those from previous studies^11-13^ do not provide clear evidence that sleep duration has a causal effect on HbA1c, glucose or T2D and that previous observational results^3 4^ are potentially explained by confounding.

Lastly, our findings suggested that previous observational estimates^44 45^ linking evening preference and daytime napping with hyperglycaemia are potentially influenced by confounding as we found no convincing evidence across MVR in one of the largest studies to date, 1SMR and 2SMR, and numerous sensitivity analyses. Ours is the first MR study of daytime napping with glycaemic traits. Two previous MR studies of the effect of chronotype on T2D reported differing results.^20 47^ Jones *et al* found no clear association of chronotype with T2D in a 2SMR design,^20^ whereas, Adams *et al* suggested a protective effect of morning preference on T2D via lowering circulating total fatty acids.^47^ These are broadly consistent with our findings that evening preference is associated with higher HbA1c in MVR and in our main 1SMR, but not in 1SMR sensitivity analyses or the main and sensitivity 2SMR analyses.

### Strengths and limitations

Key strengths of this study are: a) the comparison of results from different methods with differing key sources of bias;^10^ b) the range of sensitivity analyses that were performed for the MR analyses, including applying novel methods for the first time accounting for biases due to unbalanced horizontal pleiotropy, weak instruments, and winner’s curse in a one-sample study;^21^ and c) the use of large sample sizes of ∼340,000 for our MVR and 1SMR analyses and a sample of ∼46,000 for the 2SMR.

MVR analyses are susceptible to residual confounding, e.g. by diet, lifestyle, and physical activity, which are hard to measure precisely and hard to eliminate through statistical adjustment. Whilst we have adjusted for some potential confounders, we cannot exclude residual confounding, for example as a result of reporting bias in physical activity. The MVR analysis used cross-sectional data and it is possible that HbA1c levels could influence sleep perhaps through mechanisms including neuropathic pain and nocturia. We were able to explore this potential reverse causality using bidirectional 1SMR and 2SMR and found no evidence of an effect of HbA1c on insomnia (**Supplementary Table 9** and **Supplementary Method**).

Since a diabetes diagnosis and use of insulin medication will alter glycaemic levels, we performed subsequent analyses to exclude these individuals, which attenuated the estimates. The attenuation of the insomnia-HbA1c association could be because the measured HbA1c value in people with treated diabetes is lower than the genetic prediction (for lifetime HbA1c not on treatment) or because the impact of insomnia on HbA1c is greater in those with diagnosed diabetes than in the rest of the population. To explore the latter would require very large sample sizes that enabled stratified analyses and had sufficient power to detect interactions in the MR analyses. Currently, such samples are not available. In addition, we cannot rule out the possibility that excluding/restricting people with diabetes could introduce collider bias,^48^ given diabetes could be considered as a common effect of sleep traits and hyperglycaemia, though whether this bias is large enough to be of practical importance is doubtful.^49^

All sleep traits used were self-reported, which might be subject to measurement error.^50^ As participants will not know their value for the genetic IV, and are unlikely to know their HbA1c or glucose levels any such error would be random in relation to our outcomes and therefore expected to bias towards the null in MR and MVR analyses. In MR analyses, particularly in 2SMR, we would not expect this bias to be substantial.^51^ While the assessment of sleep traits on glucose levels was secondary to our primary aim, it should be highlighted that both MVR and 1SMR were conducted on non-fasting glucose in UKB. Analyses were adjusted for fasting time, but even with this adjustment differences between the 1SMR and 2SMR may be because the latter was based on fasting glucose.

For the majority of our MR analyses, the F-statistics and R^2^ suggested that weak instrument bias^16^ was unlikely to be substantial. This is supported by the consistency of results in sensitivity analyses which explored potential bias due to weak instruments. We found that some of the genetic instruments were associated with risk factors for HbA1c, which could result in violation of the MR assumptions through horizontal pleiotropy. This was particularly the case for the insomnia genetic instrument, which has also been shown to associated with some HbA1c risk factors other studies.^52 53^ These associations might reflect vertical pleiotropy, in which they are part of the causal path from insomnia to HbA1c (e.g. the allele score might associate with BMI because insomnia results in higher BMI). Such vertical pleiotropy would not bias our causal estimates. The associations might also highlight specific horizontal pleiotropic paths (e.g. the insomnia allele score influencing smoking independent of insomnia and generating a path from the genetic instrument to HbA1c that is not via insomnia, which could bias results.^54^) However, for insomnia, sensitivity analyses in both 1SMR and 2SMR suggested no direct (unbalanced) horizontal pleiotropy (**Supplementary Table 3** and **Supplementary Table 5**). Additionally, MVMR did not support BMI resulting in horizontal pleiotropy as results were consistent with the main analysis results.

Whilst we explored consistency across the three methods (MVR, 1SMR, and 2SMR) by focusing on the point estimates, we acknowledged that we had greater statistical power for the MVR and 1SMR than the 2SMR. It would be valuable to explore 2SMR analyses with larger outcome sample summary data. Furthermore, we cannot be certain whether the effects represent specific relations of any sleep trait. For example, the strongest evidence for a causal effect of insomnia on HbA1c was provided in our study, but GWAS significant SNPs for insomnia overlap with some for sleep duration.^14 46^ MVMR could theoretically provide some insights into this, in practice, but the genotypic and phenotypic correlations may make results unreliable even in very large sample sizes, and conditional instruments could be weak.^40^

Lastly, the low participation rate in the UKB^22^ study leads to a threat of selection bias^48^ to both observational and MR analyses. Different population sources were used in the 2SMR design, which may be less susceptible to selection bias, though in general, our 2SMR estimates provided limited supporting evidence for causality. Participants in all the study designs were predominantly of European ancestry. As such generalizing our findings to other ethnicities requires further validation.

### Implications and conclusions

We recognize that our results can be difficult to interpret clinically. For example, the interpretation of an HbA1c increase of 0.52 SD comparing those reporting “usually” experiencing insomnia to those reporting insomnia “sometimes, rarely/never” is not straightforward. We have undertaken some post estimation analyses to provide a more interpretable result. First, we converted the HbA1c results (from our main 1SMR analysis) back to the original units (mmol/mol). This shows the difference in mean HbA1c comparing “usually” *vs*. “sometimes, rarely/never” experiencing insomnia to be 1.08 mmol/mol. We then considered the impact of a hypothetical intervention that would result in everyone “usually” suffering insomnia having insomnia only “sometimes, rarely/never”. In the UK, such an intervention would need to be undertaken in 25 million adults between 40 and 70 years of age^55^ based on a “usually” insomnia prevalence of 28%, assuming it has a similar distribution of insomnia with the UKB cohort. **Box 1** provides more information for our rationale and details of this approach. If such an intervention was effective, then this would result 27,300 (95%CI 26,800 to 27,500) fewer people being diagnosed with diabetes (based on the population distribution of HbA1c and a threshold of 48 mmol/mol for diagnosing diabetes). These values could be an underestimate, since we limited our analyses to the age range of UKB (40 to 70 years old) when older and younger adults suffer from insomnia and are at risk of diabetes, and because the UKB cohort is healthier than the general UK population.^56^ They could also be an overestimate as current (largely behavioural) interventions would be challenging to implement in all people who have insomnia “usually” and none of them are effective in all participants.^57 58^ Basically we are not aware of anything that matches our hypothetical intervention in terms of being safe and effectively reducing frequent insomnia in everyone, but this exercise may help with the interpretation of the effect estimate magnitude.

#### Box 1 Quantifying the population-level impact of a hypothetical insomnia intervention on the prevalence of diabetes in the UK

Here we describe our methods for obtaining a population-level estimate of the effect of a hypothetical insomnia intervention on the UK prevalence of type 2 diabetes. We base the size of the treatment effect on the one-sample Mendelian randomization (1SMR) causal estimate in UK Biobank (UKB) participants. This causal estimate indicates if we were able to successfully intervene in all of those who “usually” experience insomnia so that the frequency of their symptoms becomes “sometimes” or “rarely/never” then this intervention would be predicted to lead to a 0.52 SD, (0.42 to 0.63) reduction in their HbA1c levels, which is equivalent to 1.08 (1.07 to 1.10) mmol/mol (1SD = 0.15 log mmol/mol in the UKB). We defined diabetic status as the level of HbA1c ≥ 48 mmol/mol. As such:

The proportion of diabetes in the UKB (before insomnia treatment, regardless of insomnia status):

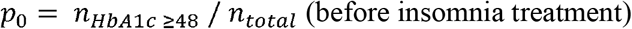

Among 28% participants in the UKB with “usually” insomnia frequency, we lower their HbA1c level by 1.08 mmol/mol. Then we re-calculated the proportion of people who have diabetes in the UKB:

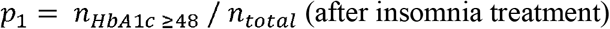

As such, the difference between *p*_O_ and *p*_l_ is the reduction in the proportion of people with diabetes as a consequence of insomnia treatment. We then used a parametric bootstrap (1,000 times) to obtain the corresponding 95% confidence interval, which is 0.109%, (0.107 to 0.110) reduction.

Lastly, we applied this proportional effect to the expected number of people in the UK between the ages of 40 and 70 years in 2018, which was the age range at baseline in UKB participants when insomnia symptoms were assessed. Approximately 38% of the UK population (i.e., 25 million out of 66 million in total) are in this age range. Therefore, 27,300 (95%CI 26,800 to 27,500) people with insomnia would be free from being diabetes in the UK.

### prevalence of diabetes in the UK

In conclusion, we presented robust evidence across MVR, 1SMR, and 2SMR studies that insomnia symptoms cause higher HbA1c. Lifestyle and/or pharmacological interventions that improve insomnia might therefore have benefits in preventing T2D. Understanding the mechanisms underlying the effect of insomnia on hyperglycaemia could help identify therapeutic strategies or new drug targets for preventing T2D. Repeating all of our 2SMR using summary GWAS from larger cohorts than those currently available would be valuable for clarifying the effects of other sleep traits on dysglycaemia.

## Data Availability

R scripts for the MVR, 1SMR, 2SMR, and other relevant sensitivity analyses are available on GitHub at: https://github.com/jamesliu0501/sleeptraits_glyacemic_MRproject.git. For statistical code relating to the individual level data analysis in UK Biobank, please contact the corresponding author via

## ACKNOWLEDGMENTS

This research has been conducted using the UK Biobank Resource under application 6818. We would like to thank the participants and researchers from the UK Biobank who contributed or collected data. Data on glycaemic traits have been contributed by MAGIC investigators and have been downloaded from www.magicinvestigators.org. We thank Dr. Eleanor Sanderson for providing statistical support on multivariable Mendelian randomization.

## Data sharing

R scripts for the MVR, 1SMR, 2SMR, and other relevant sensitivity analyses are available on GitHub at: https://github.com/jamesliu0501/sleeptraits_glyacemic_MRproject.git. For statistical code relating to the individual level data analysis in UK Biobank, please contact the corresponding author via. Summary data on glycaemic traits were downloaded from the MAGIC from www.magicinvestigators.org.

## FOOTNOTES

### Completing interest / Disclosure

All authors have completed the ICMJE uniform disclosure form at www.iemje.org/coi_disclosure.pdf and declare: MKR reports receiving research funding from Novo Nordisk, consultancy fees from Novo Nordisk and Roche Diabetes Care, and modest owning of shares in GlaxoSmithKline all unrelated to this work. JB reports receiving consultancy fees from Novartis unrelated to this work. DAL has received support from Roche Diagnostics and Medtronic Ltd for research unrelated to that presented here. No support from any organisation that might have an interest in the submitted work in the previous three years; no financial relationships with any organisation that might have an interest in the submitted work in the previous three year; no other relationship or activities that could appear to have influenced the submitted work.

### Statement

The Corresponding Author (Junxi Liu) has the right to grant on behalf of all authors and does grant on behalf of all authors, an exclusive licence (or non-exclusive for government employees) on a worldwide basis to the BMJ Publishing Group Ltd to permit this article (if accepted) to be published in BMJ editions and any other BMJPGL products and sublicences such use and exploit all subsidiary rights, as set out in our licence.

### Author contributions

MR, JB, DAL, HSD, JML, SJ, AW, TF, MC, SN, RG, SA, DR, MW and RS obtained funds for the project; RCR, JB, DAL and MR designed this study, including the developing the analysis plan; JL and RCR carried out the data analysis; CB and JB developed the novel one-sample pleiotropy robust analyses; JL, RCR, JB, DAL and MR wrote the initial draft, with subsequent input from other co-authors. All authors made critical revisions to the paper. JL, RCR and DAL act as guarantors for data integrity.

### Transparency statement

Transparency: The lead author (JL) affirms that this manuscript is an honest, accurate, and transparent account of the study being reported; that no important aspects of the study have been omitted; and that any discrepancies from the study as planned (and, if relevant, registered) have been explained.

### Ethical approval

UK Biobank has received ethical approval from the UK National Health Service’s National Research Ethics Service (ref 11/ NW/0382).

## Supplementary tables

**Supplementary Table 1**: Summary of genome-wide significant genetic instruments of sleep traits in the discovery genome-wide association studies and specific instrument strength applying to the UK Biobank and the MAGIC Consortium

**Supplementary Table 2**: Baseline characteristics of excluded participants in the UK Biobank

**Supplementary Table 3**: Multivariable regression, one-sample and two-sample Mendelian randomization analysis to assess the effects of sleep traits on HbA1c

**Supplementary Table 4:** Multivariable Mendelian randomization analysis to assess the effects of insomnia and BMI on HbA1c in the UK Biobank

**Supplementary Table 5**: Multivariable regression, one-sample and two-sample Mendelian randomization analysis to assess the effects of sleep traits on glucose

**Supplementary Table 6**: Instrument strength of the unweighted allele scores in the UK Biobank and instrument strength of individual genetic variants in the discovery genome-wide association studies and the UK Biobank

**Supplementary Table 7**: Proxies of genetic variants of sleep traits used in two-sample Mendelian randomization analysis

**Supplementary Table 8**: Associations of sleep traits unweighted allele score with risk factors for diabetes in the UK Biobank

**Supplementary Table 9**: One-sample and two-sample Mendelian randomization analysis to assess the effects of HbA1c on insomnia

