## Supplementary Methods for "Assessing the causal role of sleep traits on glycated haemoglobin: a Mendelian randomization study"

Junxi Liu, senior research associate*Ω^1, 2^, Rebecca C Richmond, research fellow*^1, 2^, Jack Bowden, professor^3, 1^, Ciarrah Barry, PhD student^1, 2^, Hassan S Dashti, research fellow^4, 5, 6^, Iyas Daghlas, MD candidate^4, 5, 6^, Jacqueline M Lane, postdoctoral researcher^4, 5, 6^, Samuel E Jones, postdoctoral researcher^7^, Andrew R Wood, lecturer^8^, Timothy M Frayling, professor^8^, Alison K Wright, research associate^9^, Matthew J Carr, research fellow^10, 11, 12^, Simon G Anderson, professor^13, 14^, Richard Emsley, professor^15^, David Ray, professor^16, 17^, Michael N Weedon, associate professor^8^, Richa Saxena, associate professor^4, 5, 6, 18^, Deborah A Lawlor, professor^1, 2, 19^, Martin K Rutter, professor^†9, 20^

* = Joint first authors, with equal contributions; ^†^ = Joint senior authors, with equal contributions; Ω Corresponding author

**Affiliations:**

1. MRC Integrative Epidemiology Unit at the University of Bristol, Bristol, UK

2. Population Health Sciences, Bristol Medical School, University of Bristol, Bristol, UK

3. College of Medicine and Health, the University of Exeter, Exeter, UK

4. Centre for Genomic Medicine, Massachusetts General Hospital, Harvard Medical School, Boston, MA, USA

5. Broad Institute of MIT and Harvard, Cambridge, MA, USA

6. Department of Anaesthesia, Critical Care and Pain Medicine, Massachusetts General Hospital, Boston, MA, USA

7. Institute for Molecular Medicine Finland, University of Helsinki, Uusimaa, Finland

8. Genetics of Complex Traits, University of Exeter Medical School, Exeter, UK

9. Division of Diabetes, Endocrinology and Gastroenterology, School of Medical Sciences, Faculty of Biology, Medicine and Health, University of Manchester, Manchester, UK

10. Division of Pharmacy and Optometry, School of Health Sciences, Faculty of Biology, Medicine and Health, University of Manchester, Manchester, UK

11. Manchester Academic Health Science Centre, University of Manchester, Manchester, UK

12. National Institute for Health Research (NIHR) Greater Manchester Patient Safety Translational Research Centre, University of Manchester, UK

13. George Alleyne Chronic Disease Research Centre, Caribbean Institute of Health Research, University of the West Indies, Kingston, Jamaica

14. Division of Cardiovascular Sciences, School of Medical Sciences, Faculty of Biology, Medicine and Health, University of Manchester, UK

15. Department of Biostatistics and Health Informatics, King’s College London, London, UK

16. National Institute for Health Research (NIHR) Oxford Biomedical Research Centre, John Radcliffe Hospital, Oxford UK

17. Oxford Centre for Diabetes, Endocrinology and Metabolism, University of Oxford, Oxford UK

18. Division of Sleep and Circadian Disorders, Brigham and Women’s Hospital, Harvard Medical School, Boston, MA, USA

19. National Institute for Health Research (NIHR) Bristol Biomedical Research Centre, University Hospitals Bristol NHS Foundation Trust and the University of Bristol, Bristol, UK

20. Diabetes, Endocrinology and Metabolism Centre, Manchester University NHS Foundation Trust, Manchester Academic Health Science Centre, Manchester, Manchester, UK

Ω Corresponding author:

Dr. Junxi Liu

MRC Integrative Epidemiology Unit, Bristol Medical School, University of Bristol

Oakfield House, Oakfield Grove, Clifton, Bristol, BS8 2BN

### UK Biobank

#### *Methods for generating the instrumental variable used in one-sample Mendelian randomization*

Biallelic and autosomal single nucleotide polymorphisms (SNPs) identified in genome-wide association study (GWAS) of self-reported sleep traits were used in the UK Biobank (UKB):^1^ 245 SNPs for insomnia,^2^ 77 SNPs for sleep duration,^3^ 27 SNPs for short sleep (≤ 6hours *vs* 7-8 hours),^3^ 7 SNPs for long sleep (≥ 9hours *vs* 7-8 hours),^3^ 37 SNPs for excessive daytime sleepiness,^4^ 114 SNPs for napping,^5^ and 341 SNPs for chronotype^6^ (specific SNPs can be checked in **Supplementary Table 6**). After the identification of genetic variants from the discovery GWAS, we recoded the SNPs in the UKB to ensure they were aligning with the discovery GWAS, in the direction of specific sleep traits’ increasing allele. To be noticed, the chronotype increasing allele was coded for morning preference (categories were ordered from more ‘eveningness’ to more ‘morningness’) in the discovery GWAS of chronotype.^6^ We have flipped the evening preference alleles for chronotype for a better interpretation (where ‘definitely a morning person’ is the reference category). Accordingly, the unweighted allele scores of each sleep traits were generated by summing the number of effect alleles harboured by each individual.

#### *Details of Self-reported sleep traits – exposures*

To assess the frequency of insomnia symptoms, participants were asked: “Do you have trouble falling asleep at night or do you wake up in the middle of the night?” with responses “Never/rarely”, “Sometimes”, “Usually”, “Prefer not to answer”, and “Do not know”. Those who responded “Prefer not to answer” or “Do not know” were set into missing. We derived a binary variable for the frequency of insomnia symptoms where “Usually” was coded as 1 and “Never/rarely” or “Sometimes” were coded as 0.

24-hour sleep duration was assessed by asking: “How many hours sleep do you get in every 24 hours? (please include naps)”. The answer could only contain integer values. Binary variables for short sleep duration (≤6 hours *vs* 7-8 hours) and long sleep duration (≥9 hours *vs* 7-8 hours) were also derived.

Self-reported daytime sleepiness was ascertained using the question “How likely are you to dose off or fall asleep during the daytime when you don’t mean to? (e.g. when working, reading or driving)” with the response options of “Never/rarely”, “Sometimes”, “Usually”, “All of the time”, “Prefer not to answer”, and “Do not know”. Participants reporting “Prefer not to answer” or “Do not know” were set into missing. Other responses were coded as 1 to 3 corresponding to the severity of daytime sleepiness.

To assess daytime napping, participants were asked: “Do you have a nap during the day?” with responses “Never/rarely”, “Sometimes”, “Usually”, “Prefer not to answer”, and “Do not know”. Those who responded “Prefer not to answer” or “Do not know” were set into missing. We derived a three-levels ordinal variable for napping frequency where “Never/rarely,” “Sometimes,” and “Usually” were coded as 1, 2, and 3, respectively.

Chronotype was assessed in the question “Do you consider yourself to be?” with the following answers: “Definitely a ‘morning’ person”, “More a ‘morning’ than an ‘evening’ person”, “Do not know”, “More an ‘evening’ than a ‘morning’ person”, “Definitely an ‘evening person”, and “Prefer not to answer” which were coded from 1 to 5 and missing respectively.

#### *Details of covariables adjusted for in multivariable-adjusted regression model*

At baseline assessment, participants completed a touchscreen questionnaire which included questions about sociodemographic status, lifestyle and environment, early life and family history, health and medical history, and psychosocial factors. Main potential confounders of the associations between sleep traits and glycaemic levels were considered to be (Model 1): age at recruitment, sex, assessment centre, smoking, alcohol intake, deprivation (Townsend residential area deprivation score^7^), university education, and vigorous physical activity levels. Body mass index (BMI) was additionally adjusted for due to uncertainty as to whether BMI was a confounder or a mediator on the causal pathway between sleep traits and glycaemic levels in Model 2.

Of the lifestyle and environment questions, participants were asked their smoking status (categorised into ‘never’, ‘former’ or ‘current’) and their alcohol intake frequency (categorised into ‘never’, ‘occasionally’, ‘1-3 times a month’ ‘once or twice a week’, ‘3-4 times a week’ or ‘daily’). Participants were also asked how many days in a typical week that they would do 10 or more minutes of vigorous physical activity (“activities that make you sweat or breathe hard such as fast cycling, aerobic exercise and heavy lifting”). Participants were asked which qualifications they had. A binary yes/no variable was generated corresponding to whether or not they reported holding a College or University degree in this question. Townsend deprivation index^7^ was calculated based on the preceding national census output areas, where each participant was assigned a continuous score corresponding to the output area in which their postcode was located. A higher index indicates a greater level of deprivation. At the initial Assessment Centre visit, height (cm) was measured using a Seca 202 device in all participants in the UKB along with sitting height while weight (kg) was measured by a variety of means, which was amalgamated into a single weight variable. BMI was calculated from height and weight in kg/m^2^.

### Methods for assessing associations of sleep traits with glucose

Glucose was measured in the same unit of mmol/l in UKB (non-fasting glucose, n = 293,838) and in Meta-Analyses of Glucose and Insulin-related traits Consortium (MAGIC) (fasting glucose, n = 46,186, mean (SD) age = 52 years (56% female) from 21 GWAS).^8^ In the UKB, non-fasting glucose was right skewed, therefore, we natural log-transformed it and converted it into SD units (1 SD = 0.17 log mmol/l). In two-sample Mendelian randomization (2SMR), we also presented results in SD units of the summary data from MAGIC (1SD was equal to an fasting glucose value of 0.73 mmol/l). Thus, for all analyses (multivariable regression (MVR), one-sample Mendelian randomization (1SMR), and 2SMR) we estimated the mean difference in glucose SD per 1 unit or category increase in the sleep traits (i.e., 24-hour sleep duration, daytime sleepiness, daytime napping, and chronotype) except for insomnia, short sleep (≤6 hours *vs* 7-8 hours), and long sleep (≥9 hours *vs* 7-8 hours). For these binary exposures, in MVR and 1SMR we estimated the average difference in glucose under the counterfactual assumption which provides an estimate of the difference between everyone (in the population of interest) experiencing the exposure (i.e. assuming exposure prevalence is 100%) compared to no-one experiencing the exposure (assuming exposure prevalence: 0%).^9^ To enable the comparison of the 2SMR estimates to the MVR and 1SMR results, we converted the results of the SNP-binary sleep trait from the multiplicative log odds scale to a difference in risk scale by $\beta=\log OR* \mu*\left( 1-\mu\right), se= {se}_{\log OR}* \mu*\left( 1-\mu\right),$ with $\mu= {n_{case}}/{(n_{case}+ n_{control})}$.^10^

In UKB, glucose was measured without fasting, because participants were not advised to fast before attending. However, participants were asked to record the last time they ate or drank anything before attending the clinic and those answers were used as ‘fasting time’. As such, we repeated MV and 1SMR main analyses with additional adjustment for fasting time (hours) and dilution factor. This dilution factor adjustment was performed because of an inadvertent dilution of serum samples which occurred during initial sample collection and processing in some samples.^11^

### One-sample and two-sample Mendelian randomization assessing associations of HbA1c with insomnia

1SMR and 2SMR were conducted to assess the association of HbA1c with insomnia to rule out the possibility of reverse causality that HbA1c levels could influence sleep perhaps through mechanisms including neuropathic pain and nocturia.

For 1SMR, 11 genome-wide significance (p-value<5×10^-8^) SNPs predicting HbA1c were identified in Soranzo N et al GWAS (n=46,368, aged 53 years-old (52% female), from 23 GWAS)^12^ from MAGIC. We generated the unweighted allele score as the total number of HbA1c increasing alleles present for each participant in the UKB. Two-stage least squares (2SLS) instrumental variable analyses were performed with adjustment for assessment centre and 40 genetic principal components to minimize confounding by population stratification, as well as baseline age, sex and genotyping chip to account for known confounders and to reduce random variation.

2SMR analyses of HbA1c with insomnia were conducted using the summary associations between the genetic instruments and HbA1c identified in the in Soranzo N et al’ s GWAS^12^ (exposure, sample 1) and estimates the associations between the genetic instruments and insomnia^2^ (downloaded from https://ctg.cncr.nl/software/summary_statistics) (outcome, sample 2). Analyses were conducted using the “TwoSampleMR” package in R.^13^ All the 11 (non-palindromic) SNPs identified in the exposure GWAS can be merged in the outcome summary statistics. Inverse-variance weighted (IVW) regression under a multiplicative random-effects model^14^ was used as the primary 2SMR analysis, meanwhile, weighted median (WM) and MR-Egger were also applied as sensitivity analyses.

### Multivariable Mendelian randomization assessing the direct effect of insomnia on HbA1c independent of BMI

97 genome-wide significance SNPs predicting BMI were extracted from Locke et al GWAS.^15^ Among which, 96 SNPs were identified and were used to generated the unweighted allele scores as the total number of BMI increasing alleles present for each participant in the UKB. We conducted a multivariable Mendelian randomization^16^ (MVMR) to assess the direct effect of insomnia on HbA1c independent of BMI in the UKB. The Sanderson-Windmeijer F statistics^16^ of insomnia unweighted allele score and BMI unweighted allele score were 1,596 and 3,970 respectively. For comparison, unique variable Mendelian randomization (UVMR) for the effect of BMI on HbA1c was also conducted separately in the UKB. Both MVMR and UVMR were performed with adjustment for age at recruitment, sex, assessment centre, 40 genetic principal components, and genotyping chip.

### Details of the novel methods used in one-sample Mendelian randomization sensitivity analyses

The method of Barry et al, termed `collider-correction’, enables weak instrument and pleiotropy robust 2SMR methods to be applied to one-sample data to obtain causal estimates.^17^ It is based on a generalization of the algorithm described in Dudridge et al , to adjust for collider bias in genetic association studies of disease progression.^18^ This method artificially induces and then corrects for collider bias, with an additional simulation extrapolation (SiMEX) correction^19^ step to cope with weak instrument bias. Further methodological details are provided in the link publication (<https://www.medrxiv.org/content/10.1101/2020.10.20.20216358v1>), but we provide a brief description below.

The association among SNP (G), exposure (X), and outcome (Y) for subject *i* is assumed to obey the following data generating process:

$X_{i} |G_{i} , U_{i}= \sum_{j=1}^{k} \beta_{XGj} G_{ij}+\beta_{UX} U_{i}+ \varepsilon_{Xi}$	(1)

$Y_{i} |X_{i} ,G_{i} , U_{i}= \beta X_{i}+\sum_{j=1}^{k} \alpha_{j} G_{ij}+\beta_{UY} U_{i}+ \varepsilon_{Yi}$	(2)

where U is the unmeasured confounding predicting X and Y, ε is the independent residual error term.

In order to obtain the unbiased causal effect $\beta$ using two stage least square (2SLS) which assuming there is no pleiotropy ($\alpha_{j}=0$), we would firstly regress the exposure (X) on all $k$ genetic variant under model (1) to derive the predicted exposure $\hat{X_{i}}$ = $\sum_{j=1}^{k} \beta_{XGj} G_{ij}$. Subsequently, we would regress the outcome (Y) on $\hat{X_{i}}$ to obtain the causal estimate $\hat{\beta}$. However, in a one-sample setting, the unmeasured confounder U is common to both X and Y, therefore the respectively residual error $\varepsilon_{Xi}$ and $\varepsilon_{Yi}$ are correlated, which might bias the estimate toward the observational estimate as long as the instruments are weak (F-statistics <10).^20 21^

Implementing collider correction

Artificially, we introduce collider bias into the SNP-outcome associations by fitting model (3):

$Y_{i} | X_{i} , G_{i}= \beta^{*} X_{i}+ \sum_{j=1}^{k} \alpha_{j}^{*} G_{ij}+ \varepsilon_{i}^{'}$	(3)

where $\beta^{*}$ and $\alpha_{j}^{*}$ are collider biased estimates distinct from $\beta$ and $\alpha_{j}$, when confounding exists between the exposure and outcome exists. The parameters $\alpha_{j}^{*}$, $\alpha_{j}$, $\beta^{*}$, and $\beta$ are linked via:

$\alpha_{j}^{*}= \alpha_{j}$ + ($\beta$ - $\beta^{*}$) $\beta_{XGj}$	(4)

To estimate $\beta$ we therefore fit a linear model to obtain an estimate $\hat{(\beta-\beta^{*})}$:

$\hat{\alpha_{j}^{*}}= \alpha_{0}$ + ($\beta$ - $\beta^{*}$) $\hat{\beta_{XGj}}$+ $\varepsilon_{i}$	(5)

As such, the causal effect is then estimated as

$\hat{\beta}$= $\hat{\beta^{*}}$+ $\hat{(\beta-\beta^{*})}$	(6)

To additionally adjust for weak instrument bias we fit linear model (5) using SiMEX^19^ or another classical measurement error correction method. This method can be used because the collider correction algorithm removes the correlation in the uncertainties of $\hat{\beta_{XGj}}$ and the $\hat{\alpha_{j}^{*}}$ .

#### Implementation of collider correction into 2SMR methods

In this study, the summary statistics for collider-correction (i.e., $\beta_{XG}$, se$\beta_{XG}$, $\beta_{YG}$, se$\beta_{YG}$, $\beta^{*}$, se$\beta^{*}$, $\alpha^{*}, {se\alpha}^{*}$) were obtained from the linear regression adjusted for age, sex, chip, assessment centre, and 40 principal components. This collider correction can be implemented to different 2SMR methods regarding different assumptions of pleiotropy. The following methods were named 1SMRsensitivity1, 1SMRsensitivity2, and 1SMRsensitivity3 respectively in the manuscript, tables, and figures.

When implementing into inverse-variance weighting (IVW), we assume the mean pleiotropy is zero and the Instrument Strength Independent of Direct Effect (InSIDE) assumption. Thus, in regression (5) we can set $\alpha_{0}$ = 0 and fit using least squares. (1SMRsensitivity1)

To account for potential pleiotropy with a non-zero mean we repeat the above procedure but allow the intercept to be estimated in (5), This is equal to performing MR-Egger regression.^22^ (1SMRsensitivity2)

To account for ‘majority valid’ pleiotropy that is nevertheless potentially in violation of the InSIDE assumption, we fit model (5) with no intercept using least-absolute deviation (LAD) regression, This is close in spirit to the weighted median (WM) approach.^23^ (1SMRsensitivity3)

Applying the combination of collider correction and 2SMR methods as a sensitivity analysis, it provides an alternative to account for both pleiotropy and weak instrument bias in a 1SMR setting, which is a less biased but more precise causal estimate comparing with the application of standard 2SMR methods.

#### Winner’s curse correction

To address this, we identified subsets of genome-wide significance SNPs (p-value<5×10^-8^) of some sleep traits in other independent GWAS that did not include UKB. For insomnia: a subset of 108 SNPs were identified in Jansen et al GWAS,^2^ when analyses were run in the 23andMe separately; for excessive daytime sleepiness: a subset of 19 SNPs were identified in Wang H et al GWAS,^4^ when analyses were ran in 337,539 unrelated individuals of European Ancestries separately; for napping: a subset of 17 SNPs were identified in Dashti HS et al GWAS,^5^ when analyses were ran in the 23andMe separately; for chronotype: a subset of 72 SNPs were identified in Jones et al GWAS,^6^ when analyses were ran in the 23andMe separately. These SNPs could be accessed in the supplementary of the specific discovery GWAS studies.^2 4-6^

We did not identify a study (other than UKB) that had undertaken genome-wide analyses of sleep duration. Replication of the 78 genome-wide significant SNPs predicting sleep duration in the UKB were conducted in the CHARGE (adult, n=47,180) and the EAGLE (childhood/adolescent, n=10,554) cohorts respectively, as well as, meta-analysis of these two cohorts with the UKB (n =446,118) were presented in the discovery GWAS.^3^ Despite, the summary statistics were driven by the UKB considering to the larger sample size of UKB. Besides, no summary statistics of meta-analysis of these two independent cohorts were given in the discovery GWAS.^3^ Although we have conducted a meta-analysis to obtain the summary statistics from the CHARGE and the EAGLE, no genome-wide significance SNP was identified. As such, no winner’s curse robust sensitivity analysis was conducted.

10. Ruth Mitchell E, BL, Mitchell, R, Raistrick, CA, Paternoster, L, Hemani, G, Gaunt, TR. MRC IEU UK Biobank GWAS pipeline version 2.

11. Biomarker assay quality procedures: approaches used to minimise systematic and random errors (and the wider epidemiological implications) UK Biobank; 2019 [Available from: <https://biobank.ctsu.ox.ac.uk/crystal/crystal/docs/biomarker_issues.pdf>.

12. Soranzo N, Sanna S, Wheeler E, et al. Common variants at 10 genomic loci influence hemoglobin A(1)(C) levels via glycemic and nonglycemic pathways. *Diabetes* 2010;59(12):3229-39. doi: 10.2337/db10-0502 [published Online First: 2010/09/23]

13. Hemani G, Zheng J, Elsworth B, et al. The MR-Base platform supports systematic causal inference across the human phenome. *Elife* 2018;7 doi: 10.7554/eLife.34408 [published Online First: 2018/05/31]

14. Burgess SB, Jack. Integrating summarized data from multiple genetic variants in Mendelian randomization: bias and coverage properties of inverse-variance weighted methods. 2015
